# Improving reproducibility of proton MRS brain thermometry: theoretical and empirical approaches

**DOI:** 10.1101/2021.10.11.21264146

**Authors:** Zhengchao Dong, Joshua T. Kantrowitz, J. John Mann

## Abstract

**Purpose:** In ^1^H MRS-based thermometry of brain, averaging temperatures measured from more than one reference peak offers several advantages including improving the reproducibility, i.e., precision, of the measurement. This paper proposes theoretically and empirically optimal weighting factors to improve the weighted average of temperatures measured from three references.

**Methods:** We first proposed concepts of equivalent noise and equivalent signal-to-noise ratio in terms of frequency measurement and a concept of relative frequency that allows the combination of different peaks in a spectrum for improving the precision of frequency measurement. Based on these, we then derived a theoretically optimal weighting factor and proposed an empirical weighting factor, both involving equivalent noise levels, for a weighted average of temperatures measured from three references, i.e., the singlets of NAA, Cr, and Ch, in ^1^H MR spectrum. We assessed these two weighting factors by comparing their errors in measurement of temperatures with the errors of temperatures measured from individual references; we also compared these two new weighting factors with two previously proposed weighting factors. These errors were defined as the standard deviations (SDs) in repeated measurements or in Monte Carlo studies.

**Results:** Both the proposed theoretical and empirical weighting factors outperformed the two previously proposed weighting factors as well as the three individual references in all phantom and in vivo experiments. In phantom experiments with 4 Hz or 10 Hz line broadening, the theoretical weighting factor outperformed the empirical one, but the latter was superior in all other repeated and Monte Carlo tests performed on phantom and in vivo data.

**Conclusion:** The proposed weighting factors are superior to the two previously proposed weighting factors and can improve the reproducibility of temperature measurement using the ^1^H MRS-based thermometry.

## 1. Introduction

^1^H MRS-based thermometry of the brain seeks to measure brain temperature and its variation across the brain.^1-5^ Unlike other MR-based thermometric methods that measure relative temperature changes spatially or temporally, and unlike other invasive techniques that enable measurement of absolute temperature with neurosurgical interventions, ^1^H MRS-based thermometry can measure absolute temperature, non-invasively.^6^ This makes this technique especially useful in pre-clinical or clinical physiology and pathophysiological studies such as those measuring temperature of neonatal brain,^7^ diagnosing intracranial tumors,^8^ monitoring brain trauma^9^ and image-guided thermal ablation.^10^ Another less exploited feature of ^1^H MRS-based thermometry is that it allows quantification of brain metabolites simultaneously, without an additional scan. ^1^H MRS can be used to study correlations between temperature and metabolism in brain development, pathophysiology of disorders and with treatment.^11, 12^

The ^1^H MRS thermometry measures temperature based on frequency differences between temperature-dependent water and temperature-independent references (e.g., prominent singlets of metabolites). A commonly used reference is the peak of N-acetyl-aspartate (NAA) at 2.01 ppm. Simultaneous use of multiple references, such as NAA, creatine (Cr), and choline (Ch) peaks, is preferrable because: (1) a preselected reference may not be sufficiently prominent^13^ or even undetectable;^14^ and (2) weighted average of temperatures derived from multiple references *may* improve reproducibility of temperature measurement.^8,15^ However, the performance of a weighted average of temperatures depends on the averaging weighting factors and an unoptimized average may be inferior to the best single reference.^15^

Here we proposed a concept of *equivalent noise* in terms of frequency measurement based on the Cramer-Rao Lower Bound (CRLB) for frequency and derived an optimized weighting and presented an empirical weighting for weighted average of temperatures obtained from multiple references. We assessed our weightings and compared their performances with other previously proposed weightings using phantom data and in vivo data from human brain.

## 2. Methods and materials

### 2.1 Theory

#### 2.1.1 Cramer-Rao lower bounds and equivalent noise for frequency measurement

We calculated the CRLB of frequency measurement for an MRS signal with Lorentzian line shape:

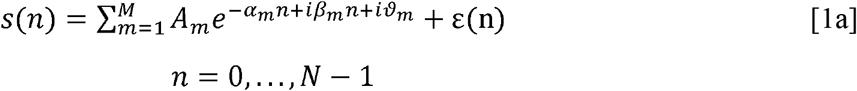

where *N* is the number of points in the time domain signal, *M* is total number of signal components, ***A***, *α, β*, and □ are constants for amplitude, normalized decay, normalized circular frequency and phase, respectively, and ε is the Gaussian noise with standard deviation (SD) □_*0*_. *α* and *β* are related to the linewidth *W*, resonance frequency *f* and spectral width *sw* by the following equations (the subscript *m* is omitted for simplicity):

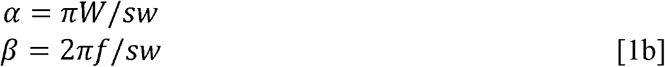

The CRLB for circular frequency is:^16^

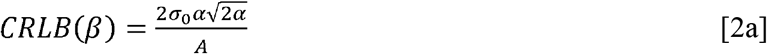

or

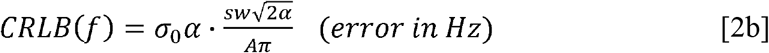

which is the preferred form for frequency in Hz and is derived from Eqs. 1b and 2a. The CRLB of frequency is the achievable minimal variance of a measurement, which is proportional to the 1.5^th^ power of the normalized decay rate and inversely proportional to the amplitude of the signal. We defined the CRLB in Eq. 2 as the equivalent noise *σ*_*e*_ in terms of frequency measurement (Figure 1):

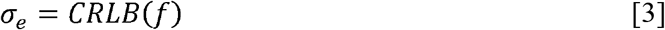

**Figure 1.**
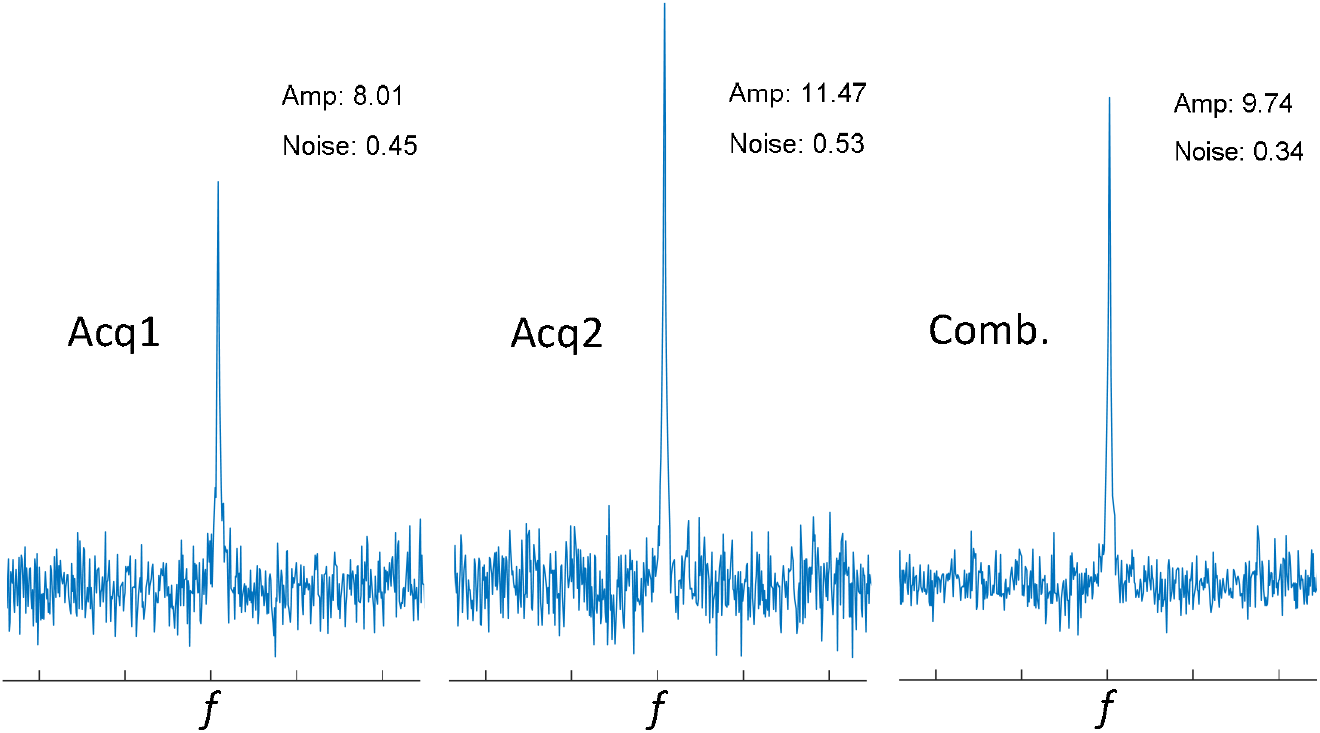
An illustration of signal averaging to enhance the SNR. Shown on the left are two acquisitions from the same sample, with different amplitudes and noise levels. After weighted averaging, the SNR of the combined signal increased (right-hand panel).

The above equation indicates that the *equivalent* noises of resonances in the same MRS spectrum may be different if their decay rates and amplitudes are different, although the spectral noise level *σ*_*0*_ is the same for all. The concept and meaning of the equivalent noise for frequency measurement are better understood from the following example.

#### 2.1.2 Improving frequency measurement by combining/averaging spectra

Signal accumulation or weighted signal averaging is a common practice in NMR for enhancing signal to noise ratio (Figure 1). When measuring the frequencies of the same peak in two spectra, the spectrum with higher *SNR*will have smaller error or give higher precision of measurement. Combining the two spectra into one and measuring its frequency, we predict, but need to verify, will increase the precision of the measurement if the *SNR* of the combined peak is greater.

We extend this logic to the averaging of multiple peaks in a *single* spectrum, by introducing the concept of relative frequencies with respect to true frequencies and employing the concept of equivalent noise (Figure 2). Here the frequencies of different peaks (both their true and measured frequencies{*f*_*i0*_} and {*f*_*i*_}^17^) are different and the peaks cannot be added. Instead, the relative frequencies {*f*_*i*_ - *f*_*i0*_}, which are the difference between the measured and the corresponding true frequencies of the individual peaks, are at the same position on the relative frequency axes, subject only to measurement errors. These peaks are therefore additive, just like the peaks in a spectrum in the above example (Figure 1). The errors of the relative frequency, measured from the combined peaks in the relative axes, can be smaller than those measured from individual peaks.

**Figure 2.**
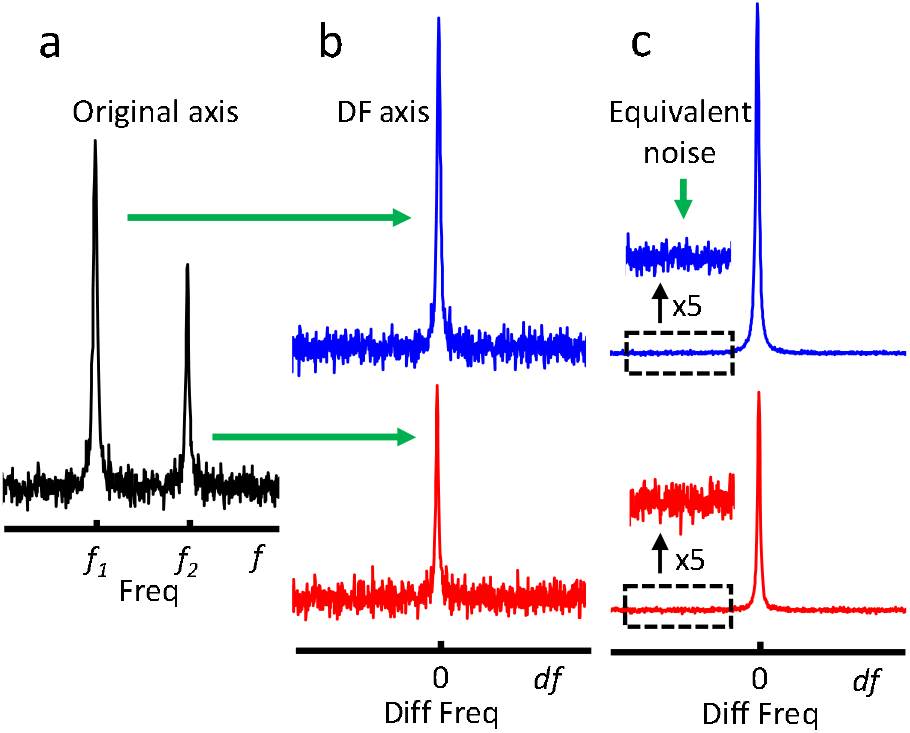
An illustration of difference frequency and equivalent noise. (a) A simulated MRS spectrum with two component peaks, each of which has its own peak height, linewidth, and resonance frequency (*f*_1_ or *f*_2_), and with Gaussian noise. (b) To facilitate the combination of the peaks at different frequencies, the two peaks are depicted on difference-frequency axis (df) with respect to their resonance frequencies of *f*_1_ and *f*_2_, respectively. (c) To account for the effect of noise on the frequency measurement, the noise levels are rescaled to the equivalent noise according to their CRLBs for frequency in Hz (Eq. 2b). Now the equivalent noise represents the achievable precision of the frequency measurement of the individual peak. Note that the df can be converted to temperature, with its origin assigned to a T_0_, eg, 37°C (see the text for details).

The conversion of the frequencies into relative frequencies has real, physical meaning in ^1^H MRST. The temperatures measured from different references are theoretically the same and are derived from the relative frequency differences of the reference with respect to water, subject to differences stemming from random noise and calibration errors. Therefore, the optimal combination of temperatures measured from references for improving precision is equivalent to optimal combination of peaks on their axes of relative frequencies. The principles of the combinations of the same peak in different acquisitions (Figure 1) and different peaks in the same acquisition (Figure 2) are the same, but the later requires that the noise levels of the individual reference peaks be converted to their equivalent noises as outlined above. The core focus of the current work is optimal averaging of frequency and eventually temperature measurement. The linear relationship between frequency and temperature, as well as between errors of frequency and temperature measurements, is given in **Appendix A**. Therefore, the terms of frequency averaging and temperature averaging are interchangeable. Furthermore, the combination of temperatures is, in essence, averaging of temperature in its technical realization, and therefore, we will mainly use “average of temperatures” in this paper.

#### 2.1.3 Weighted averaging of peaks

The weighted average of the frequencies is expressed as

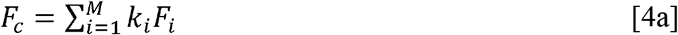

where *F*_*i*_ is frequency, *M* is the number of reference peaks, and *k*_*i*_ is the normalized weighting factor:

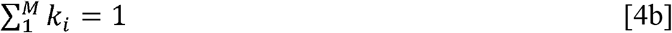

or

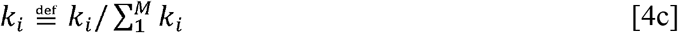

The recursive definition in Eq. 4c applies when *k*_*i*_ is not normalized. Two kinds of weighting factors before normalization were previously proposed,^15^

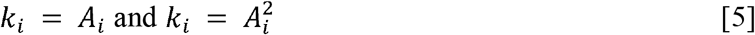

With the concept of equivalent noise, we hypothesize that an optimized weighting factor is the one that maximizes the equivalent *SNR*, or *SNR*_*e*_, of the combined peak in terms of frequency measurement.

Suppose we have an MRS signal consisting of *M* independent, exponentially decaying sinusoids as defined in Eq. 1. For simplicity, we use a 2 x *M* matrix to describe the amplitudes *A* and equivalent noises *σ*_*e*_ of the *M* component signals: (*A*_*1*_ *A*_*2*_ *… A*_*M*_; *σ*_e1_ *σ*_e2_ … *σ*_eM_). We convert the frequencies of the peaks into relative frequency axes to facilitate peak combination. Seeking an optimized weighting factor is the key to optimizing the *SNR*_*e*_ of the combined peak. The SNR of the weighted average of the peaks is expressed as follows:

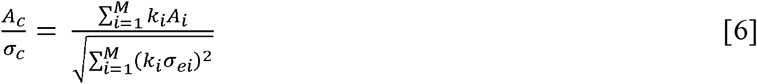

We showed theoretically that the optimal combination is realized when the weighting factor is given by (**Appendix B**):

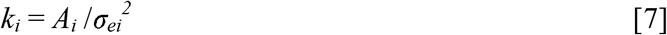

In addition to the theoretically derived weighting factor, we also propose an empirically derived weighting factor, which is the *SNR*_*e*_ itself:

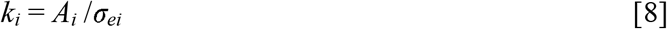

We compared and evaluated the four weighting factors (Eqs. 5 & 7, 8) using phantom and in vivo data. We used σ to represent σ_e_ in the rest of the paper for simplicity. Although we used frequencies to derive the theoretically optimal combination, Eqs. 6 - 8 hold when the *F* in Eq. 4 is replaced by the temperature *T* (**Appendix A**).

### 2.2 ^1^H MRS data acquisition and processing

#### 2.2.1 Phantom data

##### Data acquisition

We acquired single voxel phantom ^1^H MRS data on a 3T scanner (SIGNA™ Premier, GE Healthcare) using a 21-channel surface coil and PROBE-P, a commercial PRESS sequence.^18^ We used the spherical “Braino” phantom (GE Healthcare), which contains major brain metabolites with concentrations close to normal physiological values (N-acetylaspartate: 12.5mM; creatine: 10 mM; choline: 3mM; glutamate: 12.5 mM; myo-inositol: 7.5mM; lactate: 5 mM). The data acquisition parameters were as follows: TR/TE = 2000/120 ms, spectral width = 5000 Hz, FID points = 4096, number of excitations for the unsuppressed water = 16, number of saved water FIDs = 2; number of excitations for water suppressed data = 240, numbers of saved, water-suppressed FIDs (each with an 8-step phase cycling) = 30, voxel size = 4×4×4 cm^3^. Four MRS sessions were performed, each of which lasted 8 min.

##### Data preprocessing

We combined the data from coil elements using unsuppressed water signal as a reference^19,20^. We removed the residual water signal using an SVD-based method^21-23^ and performed spectral alignment among the 30 water suppressed FIDs by aligning the 2^nd^ to the 30^th^ FIDs to the 1^st^ one, using a fitting procedure like that of Near et al, using the Lorentzian lineshape function.^24^

##### Evaluation of the temperature averaging – by repeated measurements

We used two methods to evaluate the performance of temperature averaging. The first method used the repeated measurements, i.e., the 30 FIDs in each phantom MRS session. We fitted individual FIDs using a Lorentzian line shape model for the amplitudes {*A*_*i,j*_}, frequencies {*f*_*i,j*_}, decay rates {*α*_*i,j*_}, and phases {*φ*_*i,j*_}, where *i* represents NAA, Cr, and Ch, respectively, and *j* = 1 to 30 represents individual FIDs. We also calculated the original noise levels from the FIDs and converted them to the equivalent noise levels *σ*_*i,j*_, using the measured amplitudes, decay rates *α*_*i,j*_, according to Eq. 3. We converted the unit of frequencies to ppm and derived the temperatures from individual references using the calibration factors given by Zhu et al^25^. Initial results using these calibrations showed that the temperatures derived from Ch were remarkably different from those derived from NAA and Cr, and this systematic error further induced large errors mixed with random errors in the average temperatures. We therefore modified the interceptions in calibration equations so that the temperatures measured from the three references were closest for the three sets of phantom data used. The original interceptions were 313.7584, 204.6695, and 192.5210. The temperature equations with modified interceptions are as follows:

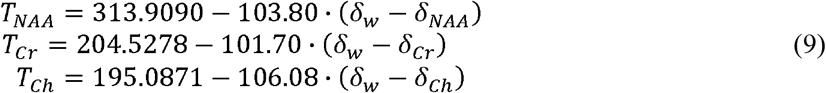

where *δ*_*w*_ is the frequency (in ppm) of the unsuppressed water signal. We measured temperatures from individual references *T*_*r,j*_ and calculated the average temperatures *T*_*c j,k*_, where *i* = {NAA, Cr, Ch}, *j* = 1, 2, …, 30, and 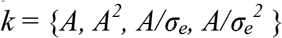. We calculated the SDs of the *T*_*r,j*_ and *T*_*c j,k*_. As an example, the latter is given as follows:

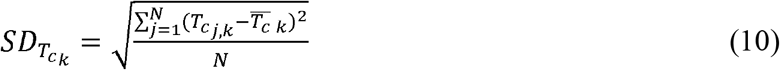

where N = 30. The optimally averaged temperatures had a mean value closest to the temperature derived from the reference with highest equivalent SNR and smallest SDs.

##### Evaluation of the temperature averaging – Effects of noise levels and linewidths

The second approach used Monte Carlo (MC) simulations, to test the effects of linewidths and noise levels, respectively, on the precision of the averaged temperatures estimated using different weighting factors. To test the effects of linewidths, we multiplied the original 30 signals in each MRS session by Lorentzian lineshape functions with linewidths of 4 and 10 Hz, respectively. We also added a complex noise signal with Gaussian distribution to individual line-broadened free induction decay (FID). The noise levels were derived from the original FIDs. We proceeded with the processing of these line-broadened and noise-added signals for the average temperatures, *T*_*c,j,k,l*_, as described above, where *l* represents the linewidth. We calculated the SDs of the averaged temperatures for each {*k, l*} and compared the effects of linewidths. The effects of noise levels were assessed following similar procedures as that for the linewidths. To test the effects of noise levels, we added noise signals with 4- and 10-times their original noise, respectively, to the 30 original signals, and proceeded to measure and calculate the SDs of the individual and averaged temperatures.

#### 2.2.2 In vivo data

##### Data acquisition

We acquired single voxel MRS data from five human subjects on the same 3T scanner as for the phantom experiments. The protocols for human studies were approved by the IRB and informed consent was obtained from each subject before the MR scan. The single voxel MRS data were acquired from medial prefrontal cortex using a 21-channel surface coil and the PROBE-P sequence^18^ with the following parameters: voxel size: 3.0 × 2.5 × 2.5 cm^3^, TR/TE = 1500/120 ms, spectral width = 2000 Hz, FID datapoints = 1024, number of saved, non-water-suppressed FIDs = 2, number of saved, water-suppressed FIDs = 30 (total number of accumulations = 240); total MRS data acquisition time was approximately 6.5 minutes.

*Data preprocessing* of the in vivo data, which included combinations of data from element coils, residual water removal and spectral alignment, was carried out using the methods as described for the phantom data. The procedures for spectral fitting and for the conversion of frequencies to temperature, also known as temperature calibration, were also performed using the methods described above. Specifically, the temperature calibration was performed for individual subjects so that the temperatures derived from the three references, NAA, Cr, and Ch, were approximately the same.

##### Evaluation of temperature averaging by Monte Carlo simulations

We used in vivo MRS datasets from each of the 5 subjects, as the basis signals in the MC simulations. We first determined the original noise level for each dataset by calculating the SDs of the data points in the second half of the FID, where the metabolite signals decayed out. We created *N* = 500 sets of complex noise signals with Gaussian distribution, and with the same noise levels as in the corresponding original, basis signals. We added these individual noises to an original, basis FID to form a set of test signals and submitted them to the MC procedure. We calculated the SDs of the averaged temperatures, *T*_*ck*_:

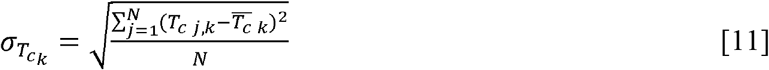

where *j* = 1, 2, …, *N* is the index of the noisy signals, *k* represents the weighting factor (Eq. 5), and 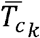 is the average of the averaged temperatures for weighting factor *k*.

## 3. Results

### Phantom experiments – original data

Examples of the spectral fitting of the phantom data, including the original, the noise added, and the line broadened spectra, are given in Figure 3. The amplitudes, frequencies, and linewidths measured for the original 30 FIDs from experiment 1 are shown in Figure 4. There was no frequency drift over the time after spectral alignment, indicating no temperature drift. There was no amplitude and linewidth variations over time, indicating stable equivalent noise levels for all three references. The reference temperatures, i.e, *T*_*NAA*_, *T*_*Cr*_, and *T*_*Ch*_, were calculated from their respective frequencies measured by spectral fitting, the average temperatures of *T*_*A*_ and *T*_*A*_^*2*^ were calculated using the amplitude-based weightings, and the average temperatures of *T*_*A/σ*_ and *T*_*A/σ*_^*2*^ were calculated using weightings from amplitudes, decays, and the noise levels. The SDs of these temperatures in the repeated measurements served as the metrics of the precisions of the temperature measurements including temperature averaging. In all four experiments (Table 1), the *A*/*σ* weighting performed best, and both *A*/*σ* and A/σ^2^ weightings outperformed the other two weightings and all three individual temperatures *T*_*NAA*_, *T*_*Cr*_ and *T*_*Ch*_. The *A*^*2*^ weighting outperformed the *A* weighting in three experiments, but it was inferior to the best individual measurement of *T*_*NAA*_.

**Figure 3.**
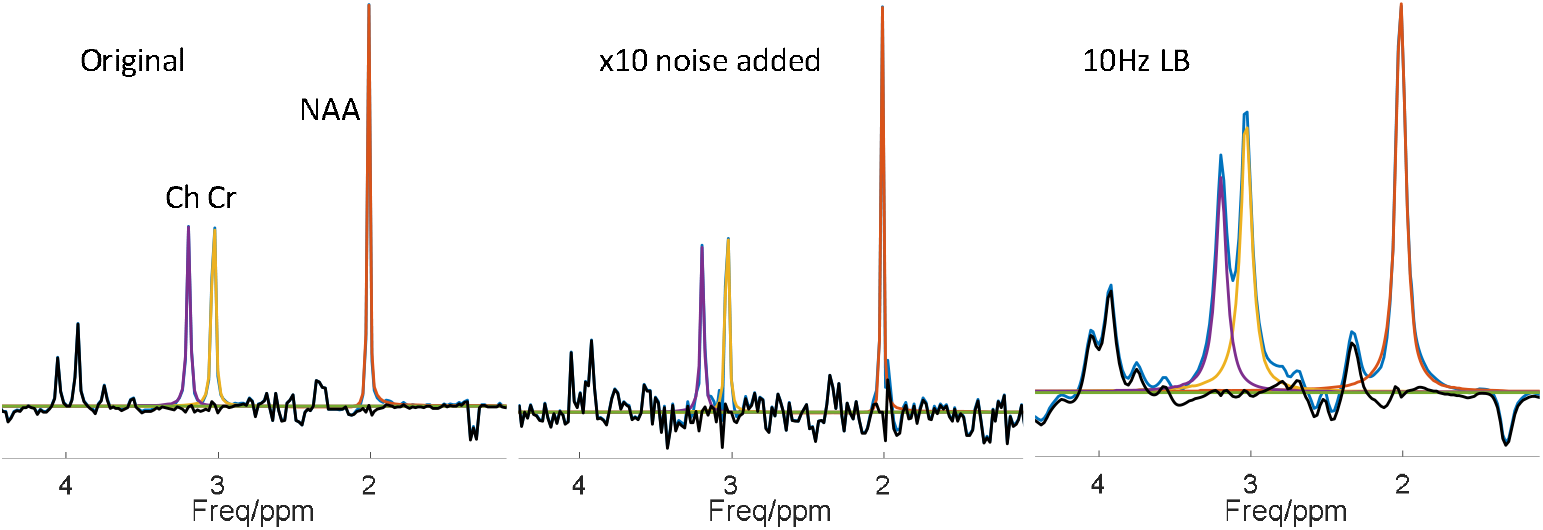
Examples of phantom spectral fitting for the original (left), x10 noise added (center), and 10 Hz line-broadened spectra (right). The spectra were fitted in the time domain using a Lorentzian line shape for NAA, Cr, and Ch. The blue and black lines are the signal to be fitted and the residue of the fitting, respectively. The other three colored lines are fitting spectra of NAA, Cr, and Ch.

**Figure 4.**
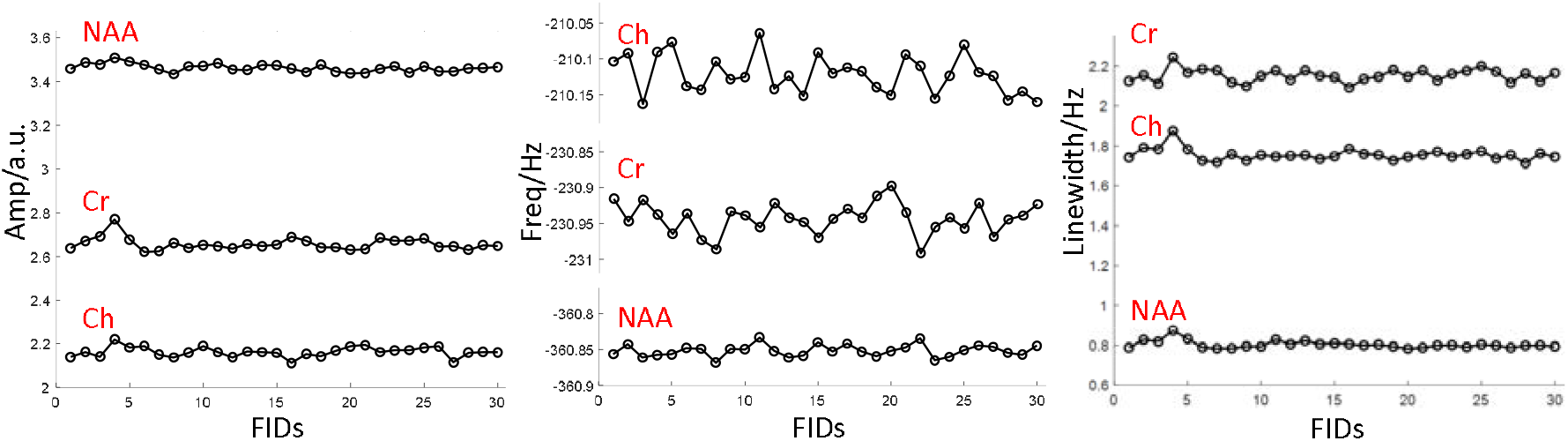
Examples of amplitudes (left), frequencies (center), and linewidths (right) of the three references measured from the 30 FIDs in an MRS session of a phantom experiment. The time interval between adjacent FIDs is 12 sec and the total time of lapse for the whole session is 6 min. Note that no frequency drift is seen over this period.

**Table 1.** Standard deviations (SDs × 10^−3^ °C) of measurements of temperatures of individual references and their combinations with different weighting factors. Data were calculated from 30 FIDs in each experiment. R2B is “Ratio to the Best”, representing the SD of a temperature to that of the best temperature measurement. The Rank is based on SDs.

| Exp. | Items | $T_{NAA}$ | $T_{Cr}$ | $T_{Ch}$ | $T_{C,A}$ | $T_{C,A}^2$ | $T_{C,A/\sigma}$ | $T_{C,A/\sigma^2}$ |
| --- | --- | --- | --- | --- | --- | --- | --- | --- |
| 1 | SD | 0.793 | 4.119 | 6.506 | 2.178 | 1.742 | 0.581 | 0.635 |
|  | R2B | 1.3649 | 7.0895 | 11.1979 | 3.7487 | 2.9983 | 1 | 1.0929 |
| 2 | SD | 2.138 | 4.888 | 9.154 | 1.973 | 2.485 | 1.803 | 1.99 |
|  | R2B | 1.1858 | 2.7110 | 5.0771 | 1.0943 | 1.3783 | 1 | 1.1037 |
| 3 | SD | 1.328 | 5.046 | 6.588 | 2.189 | 1.809 | 1.027 | 1.150 |
|  | R2B | 1.2931 | 4.9133 | 6.4148 | 2.1315 | 1.7614 | 1 | 1.1198 |
| 4 | SD | 1.4284 | 5.6820 | 7.7321 | 2.7732 | 2.2757 | 1.1484 | 1.1974 |
|  | R2B | 1.2438 | 4.9479 | 6.7332 | 2.4149 | 1.9817 | 1 | 1.0427 |

### Phantom experiments – with added noise

The results of the phantom experiments with added noise seemed trivial in that the observations about the ranking of the precisions of the temperature measurement with original data remained largely unchanged.

Overall, the *A*/*σ* weighting was still the best, but the SD of *T*_*c*_ with *A*/*σ*^*2*^ weighting was slightly lower than that of *A*/*σ* weighting for Exp. 3 with 10 times added noise (Table 2). Specifically, the SD levels of weightings *A* and *A*^*2*^ were higher than those of *A*/*σ* and *A/σ*^*2*^.

**Table 2.** Standard deviations (SDs × 10 ^-3 °^C) of temperatures of the individual references and their weighted combinations with differing weighting factors. Data were from 4 experiments, each with 30 FIDs. Noises with 4x and 10x noise levels of the original FIDs were added, respectively. R2B is “Ratio to the Best”, representing the SD of a temperature to that of the best temperature measurement.

| Noise (x) | Exp. | Items | $T_{\text{NAA}}$ | $T_{\text{Cr}}$ | $T_{\text{Ch}}$ | $T_{\text{C,A}}$ | $T_{\text{C,A}}^2$ | $T_{\text{C,A}/\sigma}$ | $T_{\text{C,A}/\sigma^2}$ |
| --- | --- | --- | --- | --- | --- | --- | --- | --- | --- |
| 4 | 1 | SD | 7.36 | 19.46 | 32.84 | 11.09 | 9.54 | 6.81 | 7.04 |
|  |  | R2B | 1.08 | 2.86 | 4.82 | 1.62 | 1.40 | 1 | 1.03 |
|  | 2 | SD | 6.00 | 11.17 | 20.41 | 7.20 | 6.30 | 5.30 | 5.72 |
|  |  | R2B | 1.13 | 2.11 | 3.85 | 1.36 | 1.19 | 1 | 1.08 |
|  | 3 | SD | 9.22 | 25.48 | 41.90 | 12.83 | 10.92 | 8.10 | 8.60 |
|  |  | R2B | 1.14 | 3.15 | 5.17 | 1.58 | 1.35 | 1 | 1.06 |
|  | 4 | SD | 8.12 | 23.68 | 36.57 | 12.24 | 10.57 | 7.72 | 7.72 |
|  |  | R2B | 1.05 | 2.07 | 4.73 | 1.59 | 1.37 | 1 | 1.00 |
| 10 | 1 | SD | 22.69 | 56.36 | 86.61 | 31.04 | 27.36 | 20.91 | 21.79 |
|  |  | R2B | 1.09 | 2.70 | 4.14 | 1.48 | 1.31 | 1 | 1.04 |
|  | 2 | SD | 20.11 | 55.38 | 88.44 | 29.55 | 25.63 | 18.40 | 19.10 |
|  |  | R2B | 1.09 | 3.01 | 4.81 | 1.61 | 1.39 | 1 | 1.04 |
|  | 3 | SD | 21.13 | 57.99 | 107.95 | 36.36 | 30.84 | 19.81 | 19.51 |
|  |  | R2B | 1.08 | 2.97 | 5.53 | 1.86 | 1.58 | 1.02 | 1 |
|  | 4 | SD | 22.35 | 60.44 | 93.85 | 30.36 | 26.40 | 20.51 | 21.16 |
|  |  | R2B | 1.09 | 2.95 | 4.58 | 1.48 | 1.29 | 1 | 1.03 |

### Phantom experiments – with line broadening

Line broadening had a remarkable influence on the precision of the averaged temperatures (Table 3). In all cases except Exp. 1 with 4 Hz line broadening, *A/σ*^*2*^ outperformed *A*/*σ* and became the best weighting factor. But the *A*/*σ* weighting was still better than the other two weightings (*A* and *A*^*2*^). The *A*^*2*^ weighting remained superior to the *A* weighting and surpassed the best individual temperature measurement *T*_*NAA*_.

**Table 3.** Standard deviations (SDs × 10^−3^ °C) of temperatures of the individual references and their weighted combinations with differing weighting factors. Data were from 4 experiments, each with 30 FIDs. The original FIDs were line broadened by 4 Hz and 10 Hz, respectively, for this test. To keep the reasonable noise levels for the data, Gaussian noises with standard deviations equal to the noise levels of the corresponding original FIDs were added to the FIDs after line broadening. R2B is “Ratio to the Best”, representing the SD of a temperature to that of the best temperature measurement.

| LB (Hz) | Exp. | Items | $T_{\text{NAA}}$ | $T_{\text{Cr}}$ | $T_{\text{Ch}}$ | $T_{\text{C,A}}$ | $T_{\text{C,A}}^2$ | $T_{\text{C,A}/\sigma}$ | $T_{\text{C,A}/\sigma^2}$ |
| --- | --- | --- | --- | --- | --- | --- | --- | --- | --- |
| 4 | 1 | SD | 8.41 | 14.55 | 20.69 | 8.26 | 7.69 | 7.15 | 7.24 |
|  |  | R2B | 1.18 | 2.03 | 2.89 | 1.16 | 1.08 | 1 | 1.01 |
|  | 2 | SD | 8.29 | 16.30 | 26.10 | 8.70 | 7.58 | 6.74 | 6.68 |
|  |  | R2B | 1.24 | 2.44 | 3.91 | 1.30 | 1.13 | 1.01 | 1 |
|  | 3 | SD | 17.80 | 32.63 | 39.66 | 16.60 | 15.46 | 14.62 | 14.42 |
|  |  | R2B | 1.23 | 2.26 | 2.75 | 1.15 | 1.07 | 1.01 | 1 |
|  | 4 | SD | 18.01 | 33.49 | 44.35 | 16.92 | 15.43 | 14.33 | 14.06 |
|  |  | R2B | 1.28 | 2.38 | 3.15 | 1.20 | 1.10 | 1.02 | 1 |
| 10 | 1 | SD | 41.81 | 67.48 | 94.09 | 35.39 | 33.09 | 31.78 | 31.72 |
|  |  | R2B | 1.31 | 2.12 | 2.97 | 1.12 | 1.04 | 1.00 | 1 |
|  | 2 | SD | 46.91 | 94.47 | 124.75 | 47.50 | 44.35 | 42.28 | 40.18 |
|  |  | R2B | 1.17 | 2.35 | 3.10 | 1.18 | 1.10 | 1.05 | 1 |
|  | 3 | SD | 55.67 | 96.01 | 134.86 | 52.90 | 48.95 | 47.30 | 45.46 |
|  |  | R2B | 1.22 | 2.11 | 2.97 | 1.16 | 1.08 | 1.04 | 1 |
|  | 4 | SD | 59.20 | 88.44 | 131.71 | 52.56 | 48.87 | 47.12 | 45.88 |
|  |  | R2B | 1.29 | 1.93 | 2.87 | 1.15 | 1.07 | 1.03 | 1 |

### In vivo experiments

The spectral fitting of the in vivo data focused on the major signals of NAA, Cr and Ch. Therefore, the J-coupled spectral peaks remained in the residue, but they did not affect the quality of the spectral fitting (Figure 5). For the data from the five subjects, the *A*/*σ* weighting outperformed other weighting factors in terms of the lowest SDs. The *A*/σ^2^, *A*, and *A*^*2*^ weightings had three, two, and one 2^nd^-place rankings, respectively, but *A* had three fourth-place rankings (Table 4). The SDs of all averaged temperatures were smaller than that of the best individual reference, which was NAA in this case, showing that all weighted averaging of temperatures improved the accuracy of temperature measurement. Overall, the proposed *A*/*σ* and *A*/σ^2^ weightings was superior to the other previously proposed weightings. The improvement of reproducibility (precision) of the optimized A/*σ*_e_-weighting over the A^2^-weighting was 3.85% ± 0.96%. The improvement of A/σ_e_-weighting over the best single reference (NAA) was 73.1% ± 21.6%.

**Table 4.** Comparison of the accuracies of temperatures derived from individual references (NAA, Cr, and Ch) and different temperature combination algorithms (with weighting factors of A, A^2^, A/σ, and A/σ^2^). The original single voxel ^1^H MRS were from five human subjects. Each MRS data set was combined with 1000 individual noise signals whose distribution is Gaussian and standard deviations are the same as those of the original signals. The above-mentioned individual temperatures from three references and averaged temperatures from four weighting factors were calculated and their standard deviations (SD ×10^−2^ °C) were presented here as a matric of the accuracy of the temperature measurement. R2B is “Ratio to the Best”, representing the SD of a temperature to that of the best temperature measurement. The Rank is based on SDs.

| Subj. | Items | $T_{NAA}$ | $T_{Cr}$ | $T_{Ch}$ | $T_{C,A}$ | $T_{C,A}^2$ | $T_{C,A/\sigma}$ | $T_{C,A/\sigma^2}$ |
| --- | --- | --- | --- | --- | --- | --- | --- | --- |
| 1 | SD | 8.2699 | 12.1774 | 8.7685 | 4.5802 | 4.5754 | 4.3746 | 4.5044 |
|  | R2B | 1.8905 | 2.7837 | 2.0044 | 1.0470 | 1.0459 | 1 | 1.0297 |
|  | Rank | 5 | 7 | 6 | 4 | 3 | 1 | 2 |
| 2 | SD | 8.2463 | 12.2642 | 9.2228 | 4.4705 | 4.4629 | 4.2816 | 4.4625 |
|  | R2B | 1.9260 | 2.8644 | 2.1540 | 1.0441 | 1.0424 | 1 | 1.0423 |
|  | Rank | 5 | 7 | 6 | 4 | 2 | 1 | 2 |
| 3 | SD | 5.4464 | 8.6015 | 6.6135 | 3.3377 | 3.3332 | 3.2381 | 3.2729 |
|  | R2B | 1.6820 | 2.6564 | 2.0424 | 1.0308 | 1.0294 | 1 | 1.0108 |
|  | Rank | 5 | 7 | 6 | 4 | 3 | 1 | 2 |
| 4 | SD | 10.1903 | 15.5803 | 13.2925 | 5.8028 | 5.9128 | 5.7561 | 6.0852 |
|  | R2B | 1.7703 | 2.7067 | 2.3093 | 1.0081 | 1.0272 | 1 | 1.0572 |
|  | Rank | 5 | 7 | 6 | 2 | 3 | 1 | 4 |
| 5 | SD | 5.3373 | 8.8891 | 6.3851 | 3.9434 | 4.0351 | 3.8502 | 4.0350 |
|  | R2B | 1.3862 | 2.3087 | 1.6584 | 1.0242 | 1.0480 | 1 | 1.0480 |
|  | Rank | 5 | 7 | 6 | 2 | 3 | 1 | 3 |

**Figure 5.**
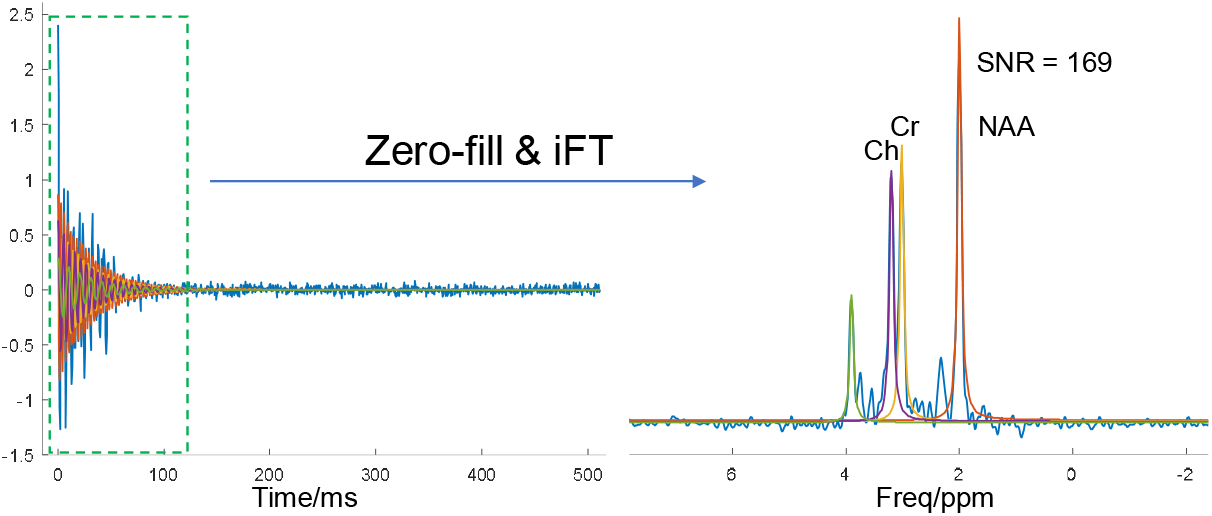
An example of spectral fitting of in vivo data. Fitting was performed in the time domain. The blue line is the signal to be fitted, and the color lines are the fitting signal components. Prior to FT, the FID was apodized and zerofilled to suppress noise in the signal. Please note that Gaussian noise with the same level of noise in the original signal was added to the spectrum to facilitate the Monte Carlo study.

## Discussion

We have proposed concepts of equivalent noise and equivalent signal to noise ratio in terms of the measurement of frequency according to the CRLBs and a concept of relative frequency. Based on these concepts, we derived a theoretical weighting factor, *A*/σ^2^, where σ is the equivalent noise, for the combination of relative frequencies of multiple references that may improve precision of temperature measurement. We also proposed an empirical weighting factor *A*/σ, which is the equivalent SNR. We carried out phantom and in vivo experiments to evaluate the performances of the two weighting factors and compared them with two previously proposed weighting factors.^15^ The results of phantom and in vivo experiments showed superiority of our two proposed weighting factors over the previously proposed ones in terms of the precision of temperature measurement.

The concept of the equivalent noise, which involves not only the conventional spectral noise, but also the amplitude and decay rate (i.e., linewidth in the frequency domain), plays a fundamental role in the development of the two proposed weighting factors. First, it makes the two weighing factors peak specific. The conventional spectral noise is global, meaning that it is the same for all components (peaks) in an MRS spectrum. Therefore, replacing the equivalent noise with the conventional, global spectral noise in the currently proposed weighting factors is meaningless, as it will be cancelled out in the normalization. This is the reason that only *A* and *A*^*2*^ weightings were previously suggested without involving the noise level.^15^ Second, the equivalent noise is directly related to the measurement precision of frequency – a larger equivalent noise means lower precision of frequency measurement. Therefore, it is intuitive to place the equivalent noise or its square into the denominator of a weighting factor: thus, a peak with larger equivalent noise should have smaller weighting in the combination, and *vice versa*.

The concept of relative frequency also played an important role in the development of the two proposed weighting factors. This concept makes different peaks in a spectrum equivalent or at the same location in the relative frequency axis, thus making possible the combination of peaks with different frequencies in a spectrum. The physical basis of the relative frequency in this paper is that the frequencies of different reference peaks correspond to the same temperature in ^1^H MRS-based thermometry. In this sense, these frequencies are equivalent, and this is reflected in the relative frequency axis.

*A*/σ^2^ weighting outperformed other weightings only in the phantom experiments with line broadening, and the results of both other phantom experiments (original data and noise-added data) and in vivo experiments showed that the empirical *A*/σ, instead of the theoretically optimal *A*/σ^2^, is the optimal weighting factor. We note that the derivation of the optimal weighting *A*/σ^2^ (**Appendix B**) is based on the optimal conditions that both the amplitudes and the decay rates did not have errors. In this case, the weighting factors can be correctly calculated. In real world data, amplitudes and decay rates have measurement errors (Eqs. 3 and 6). These errors may accumulate and propagate to affect the equivalent noise used in the proposed weighting factor, causing the weighting factor to deviate from its correct value. The error of the individual temperature is directly associated with error of the equivalent noise (Eq. 3), whose error will also in turn spoil the performance of the proposed weighting factors. *A*/σ^2^ weighting performed best when the given, noiseless amplitudes and decay rates (linewidths) were used in calculating the weighting factors. It may be inferior to *A*/σ weighting because of its larger errors in σ^2^ than in σ (∼ α^1.5^/A). This also explains why *A*^*2*^ weighting is better than *A*^*2*^ weighting in some cases, because the *Δk* error in *A*^*2*^ is double of that in *A*.

The optimal averaging of temperatures is similar to the combination of MRS data acquired using multichannel coil arrays, where equal weighting^26^, amplitude weighting (A),^27, 28^ signal to noise (*A/N*) weighting,^20^ and signal to squared noise (*A/N*^*2*^) weighting^26^ have been proposed. While the *A/N*^*2*^ weighting was theoretically derived as the optimal weighting factor, a study^21^ showed that its performance may be inferior to the *S/N* weighting in some cases. In fact, the situation here is more complex than the combination of the multichannel coil MRS data. In the latter, only amplitudes of water signal and spectral noise levels are needed, and both can be measured more accurately and precisely than the amplitudes of the reference peaks and equivalent noise levels. In the present case, not only the spectral noises and amplitudes of the reference peaks but also the decay rates (linewidths) are needed in the weighting factor, which increases the complexity and noise accumulation. Because the *A*/σ ^2^ involves the square of the equivalent noise σ, its errors caused by the errors in amplitudes, decay rates, and spectral noises would be larger than errors in *A*/σ. Therefore, the *A*/σ^2^ weighting is prone to be inferior to the *A*/*σ* weighting, more so than is the case for multichannel coil data combination.

A simple or suboptimally weighted averaging of frequencies/temperatures may not ensure improved precision, i.e., the precision of the average temperatures may not surpass the precision of the temperature measured from the best individual reference. In the experiments using original phantom data, the SDs of both *A* and *A*^*2*^ weightings were inferior to that of NAA reference (Tables 1 & 2). This can be seen in a previous study,^15^ where the SDs of all averaged temperatures using *A*^*2*^ weighting were larger than that of *T*_*Ch*_, which was derived from the dominant peak of Ch, albeit they were smaller than those of single referenced *T*_*NAA*_ and *T*_*Cr*_. Recently, Maudsley et al also found no improvement using *A*^*2*^ weighting compared with the best individual reference.^1^

The reproducibility (precision) of the temperature measurements in this paper was very high, due to the high quality of the data. In the phantom data, the average amplitude-to-noise ratios (or SNR) were > 5 times, and the decay rates (linewidths) were < 1/10 of those encountered in routine in vivo studies (**Supplementary Materials**, sTable 1). Based on the spectral fitting results of the amplitudes, decay rates, and noise levels, the CRLBs of frequencies and corresponding temperatures were in the order of 10^−3^ °C, which agreed with the measured SDs in the repeated measurements (Table 1 & sTable 1). When using realistic amplitude-to-noise ratios and decay rates, the CRLBs will be > 0.2 °C, which are close to those SD values reported in the literature (sTable 2).^9, 15^ For in vivo data, the average amplitude-to-noise ratios were > 5 times of those in routine human studies (Figure 5 & sTable 3). Therefore, the CRLBs calculated from fitting parameters agreed well with the SDs measured in the MC studies (Table 4 & sTable 3), but they were only about 1/5 of the errors in routine MRST studies.^9, 15^ The high reproducibility of the ^1^H MRST owes much to the hardware development such as the high sensitivity multi-channel coil. Independent to the hardware development, the over 73% relative improvement of the reproducibility of the optimized weighting with respect to the best single reference is remarkable.

Some technical notes are necessary. (1) We focused on improving the reproducibility or reducing random errors of the ^1^H MRS-based thermometry by optimally combining temperatures measured from three references. In ^1^H MRS-based thermometry, systematic errors or consistent biases may result from several methodological aspects of the measurement such as gradient heating,^29, 30^ separate measurement of water and reference signals,^30^ errors in the calibration parameters,^25, 31^ etc. For example, the range of frequency shift of the water suppressed spectrum may be from 0 to 9 Hz in 13 minutes with respect to the separately measured non-water suppressed spectrum, corresponding temperature errors from 0 to -3.0 °C.^30^ The present work was not dedicated to improving the accuracy or reducing the systematic errors. Therefore, we did not provide the temperature values for phantom and in vivo experiments. However, we have mitigated systematic errors in our phantom data processing by aligning the water-suppressed FIDs and avoided the mingling of systematic errors and random errors. The Monte Carlo approach to using in vivo data also avoided effects of systematic errors, where an MRS data was added with noise sets repeatedly in a Monte Carlo study. (2) The expression of the equivalent noise (Eq. 3), i.e., the CRLB for frequency, is derived and valid for singlets with Lorentzian lineshape and without overlapping with other peaks. The peaks of NAA, Cr, and Cho satisfy these conditions when the spectrum is of good quality. When the lineshape deviates from Lorentzian or peaks of Cr and Ch overlap, the equivalent noise in Eq. 3 will only be approximately correct.^9, 15, 32^ In the latter case, the actual equivalent noises of Cr and Ch will be larger than that given by Eq. 3. This explains the results in Table 4, where the SDs of Cr and Chare much larger than those of NAA. (3) We and many others^8, 15, 33, 34^ used Lorentzian lineshape to fit the spectrum. Lorentzian is the intrinsic lineshape and a commonly used model to approximate real world MR spectrum lineshapes with symmetric or asymmetric distortions. Major sources of asymmetric lineshape distortion are high order B_0_ field inhomogeneity and eddy current effect. Fitting asymmetric peaks with symmetric analytic lineshape models may in general induce systematic errors in frequency measurement. However, the systematic errors in water frequency and reference frequencies can be largely cancelled in temperature measurement when frequency difference is used in the calibration. High order temperature distribution within the MRS voxel will also cause lineshape distortion in the water peak, and this may induce systematic error in temperature measurement. However, the systematic errors were cancelled in our repeated phantom FIDs and in our in vivo data in the Monte Carlo studies. In either case, the systematic errors remained the same and did not enter the calculation of SD, which is a metric of the random error.

## Conclusion

We proposed concepts of equivalent noise, equivalent SNR, and relative frequency in terms of frequency measurement and the combination of peaks of different frequencies. Based on these concepts, we derived a theoretically optimized weighting factor and proposed an empirical weighting factor for the averaging of temperatures measured from three references. Experiments using phantom and in vivo data showed that these two weightings outperformed previously proposed weightings in improving the reproducibility of temperature measurement using the ^1^H MRS-based thermometry.

## Data Availability

Data is available subject to the regulations of the New York State Psychiatric Institute and Columbia University.No

## Abbreviations and syllables

A: amplitude
Ch: choline
Cr: creatine
CRLB: Cramer-Rao Lower Bound
MC: Monte Carlo
MRS: magnetic resonance spectroscopy
NAA: N-acetyl-aspartate
NMR: nuclear magnetic resonance
ppm: part per million
R2B: ratio to the best
SD: standard deviation
SNR: signal to noise ratio
SNR_e_: *equivalent* signal to noise ratio
T_A/σ_: weighted average temperature using weighting factor A/σ
T_c_: combined or average temperature
T_NAA_: temperature measured using NAA as a reference
δ: frequency in ppm
σ_e_: equivalent noise

## Appendix A: Temperature error due to frequency error of the reference

The temperature measured from the frequencies of water and the reference is given as follows:

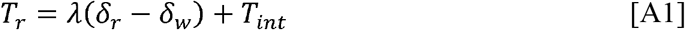

where *T*_*r*_ is the temperature measured from a reference, eg, the singlet of NAA at 2.01 ppm, *λ* is the frequency-to-temperature coefficient, *δ*_*r*_ and *δ*_*w*_ are the frequencies of the reference and water, respectively, and *T*_*int*_, is the intercept. Both *λ* and *T*_*int*_, are constants determined by the calibration experiment.

The error of *T*_*r*_ is:

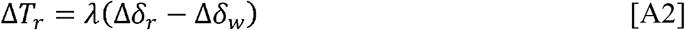

According to the CRLB (Eq. 2), the measurement error of water frequency Δ*δ*_*w*_is more than 3 order of magnitude smaller than of the reference Δ*δ*_*r*_ and, therefore, can be ignored. This results in:

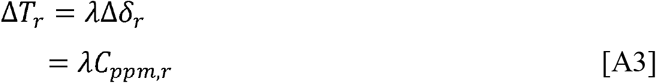

where Δ*δ*_*r*_ is substituted by the Cramer-Rao low bound for frequency measurement, *C*_*ppm,r*_.

## Appendix B: Show that 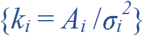 are the optimized weighting factors for the weighted averaging of multiple frequency measurements

We start from an intuitive example of two peaks with (A; σ) = (40; 10) and (10; 1), respectively. First, we let {*k*_*i*_ = *A*_*i*_}, which means the weighting factors are proportional to their corresponding amplitudes. The combined peak is (34; 8.0), whose *SNR* is *R* = 4.25 – larger than the first but small than the second peak. The reason for the failure of this weighting is that it does not take the noise into account.

Now we let {*k*_*i*_ = *A*_*i*_ /*σ*_*i*_}, meaning that the weighting factors are proportional to their {*R*}. Using the above example, we obtained the combined peak of (18.57; 2.95), whose *R* is 6.31 but is still smaller than the second one. The reason for the failure is that the SNR is not normalized but proportional to the noise. To overcome the problem, we use noise level to normalize SNR and let 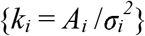. The combined peak is given by (11.15; 1.04), whose *R* is 10.77. To derive an optimized weighting, we assume 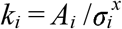 and determine an optimal *x*.

Substituting 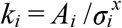into Eq. 4, we have:

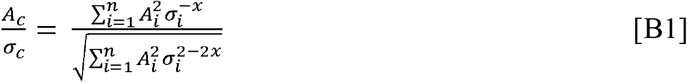

where *A*_*i*_ and *σ*_*i*_ are amplitude and noise level of the *i*-th peak, respectively.

Taking the derivative of the above equation with respect to *x*, we obtain:

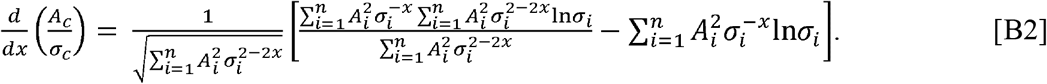

Solving

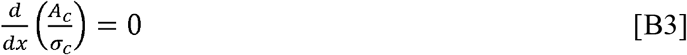

gives *x* = 2 and the optimized frequency measurement is

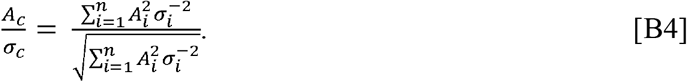

For the special case where all measurements have the same *A*_*i*_/*σ*_i_, the above equation reduces to its well-known form for signal accumulation:

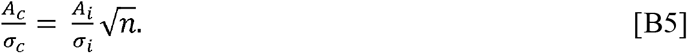

## Reference

1. Maudsley AA, Goryawala MZ, Sheriff S. Effects of tissue susceptibility on brain temperature mapping. Neuroimage 2017; 146: 1093–1101.

2. Wang P. Evaluation of MR thermometry with proton resonance frequency method at 7T. Quant Imaging Med Surg 2017; 7(2): 259–266.

3. Dehkharghani S, Mao H, Howell L, Zhang X, Pate KS, Magrath PR et al. Proton resonance frequency chemical shift thermometry: experimental design and validation toward high-resolution noninvasive temperature monitoring and in vivo experience in a nonhuman primate model of acute ischemic stroke. AJNR Am J Neuroradiol 2015; 36(6): 1128–1135.

4. Thrippleton MJ, Parikh J, Harris BA, Hammer SJ, Semple SI, Andrews PJ et al. Reliability of MRSI brain temperature mapping at 1.5 and 3 T. NMR Biomed 2014; 27(2): 183–190.

5. Sharma AA, Nenert R, Mueller C, Maudsley AA, Younger JW, Szaflarski JP. Repeatability and Reproducibility of in-vivo Brain Temperature Measurements. Front Hum Neurosci 2020; 14: 598435.

6. Odeen H, Parker DL. Magnetic resonance thermometry and its biological applications -Physical principles and practical considerations. Prog Nucl Magn Reson Spectrosc 2019; 110: 34–61.

7. Bainbridge A, Kendall GS, De Vita E, Hagmann C, Kapetanakis A, Cady EB et al. Regional neonatal brain absolute thermometry by 1H MRS. NMR Biomed 2013; 26(4): 416–423.

8. Babourina-Brooks B, Wilson M, Arvanitis TN, Peet AC, Davies NP. MRS water resonance frequency in childhood brain tumours: a novel potential biomarker of temperature and tumour environment. NMR Biomed 2014; 27(10): 1222–1229.

9. Marshall I, Karaszewski B, Wardlaw JM, Cvoro V, Wartolowska K, Armitage PA et al. Measurement of regional brain temperature using proton spectroscopic imaging: validation and application to acute ischemic stroke. Magn Reson Imaging 2006; 24(6): 699–706.

10. Zhu M, Sun Z, Ng CK. Image-guided thermal ablation with MR-based thermometry. Quant Imaging Med Surg 2017; 7(3): 356–368.

11. Posporelis S, Coughlin JM, Marsman A, Pradhan S, Tanaka T, Wang H et al. Decoupling of Brain Temperature and Glutamate in Recent Onset of Schizophrenia: A 7T Proton Magnetic Resonance Spectroscopy Study. Biol Psychiatry Cogn Neurosci Neuroimaging 2018; 3(3): 248–254.

12. Kohut SJ, Kaufman MJ. Magnetic resonance spectroscopy studies of substance use disorders: Current landscape and potential future directions. Pharmacol Biochem Behav 2021; 200: 173090.

13. Xu D, Vigneron D. Magnetic resonance spectroscopy imaging of the newborn brain--a technical review. Semin Perinatol 2010; 34(1): 20–27.

14. Horska A, Barker PB. Imaging of brain tumors: MR spectroscopy and metabolic imaging. Neuroimaging Clin N Am 2010; 20(3): 293–310.

15. Cady EB, Penrice J, Robertson NJ. Improved reproducibility of MRS regional brain thermometry by ‘amplitude-weighted combination’. NMR Biomed 2011; 24(7): 865–872.

16. Cavassila S, Deval S, Huegen C, van Ormondt D, Graveron-Demilly D. Cramer-Rao bounds: an evaluation tool for quantitation. NMR Biomed 2001; 14(4): 278–283.

17. Hartmann J, Gellermann J, Brandt T, Schmidt M, Pyatykh S, Hesser J et al. Optimization of Single Voxel MR Spectroscopy Sequence Parameters and Data Analysis Methods for Thermometry in Deep Hyperthermia Treatments. Technol Cancer Res Treat 2017; 16(4): 470–481.

18. Bottomley PA. Spatial localization in NMR spectroscopy in vivo. Ann N Y Acad Sci 1987; 508: 333–348.

19. Lin Y-Y, Hodgkinson P, Ernst M, Pines A. A Novel Detection – Estimation Scheme for Noisy NMR Signals:Applications to Delayed Acquisition Data. JOURNAL OF MAGNETIC RESONANCE 1997; 128: 30–41.

20. Dong Z, Peterson B. The rapid and automatic combination of proton MRSI data using multi-channel coils without water suppression. Magn Reson Imaging 2007; 25(8): 1148–1154.

21. Dong Z, Dreher W, Leibfritz D. Toward quantitative short-echo-time in vivo proton MR spectroscopy without water suppression. Magn Reson Med 2006; 55(6): 1441–1446.

22. Kantrowitz JT, Dong Z, Milak MS, Rashid R, Kegeles LS, Javitt DC et al. Ventromedial prefrontal cortex/anterior cingulate cortex Glx, glutamate, and GABA levels in medication-free major depressive disorder. Transl Psychiatry 2021; 11(1): 419.

23. Dong Z, Grunebaum MF, Lan MJ, Wagner V, Choo TH, Milak MS et al. Relationship of Brain Glutamate Response to D-Cycloserine and Lurasidone to Antidepressant Response in Bipolar Depression: A Pilot Study. Front Psychiatry 2021; 12: 653026.

24. Near J, Edden R, Evans CJ, Paquin R, Harris A, Jezzard P. Frequency and phase drift correction of magnetic resonance spectroscopy data by spectral registration in the time domain. Magn Reson Med 2015; 73(1): 44–50.

25. Zhu M, Bashir A, Ackerman JJ, Yablonskiy DA. Improved calibration technique for in vivo proton MRS thermometry for brain temperature measurement. Magn Reson Med 2008; 60(3): 536–541.

26. Hall EL, Stephenson MC, Price D, Morris PG. Methodology for improved detection of low concentration metabolites in MRS: optimised combination of signals from multi-element coil arrays. Neuroimage 2014; 86: 35–42.

27. Natt O, Bezkorovaynyy V, Michaelis T, Frahm J. Use of phased array coils for a determination of absolute metabolite concentrations. Magn Reson Med 2005; 53(1): 3–8.

28. Wijtenburg SA, Knight-Scott J. Reconstructing very short TE phase rotation spectral data collected with multichannel phased-array coils at 3 T. Magn Reson Imaging 2011; 29(7): 937–942.

29. Hui SCN, Mikkelsen M, Zollner HJ, Ahluwalia V, Alcauter S, Baltusis L et al. Frequency drift in MR spectroscopy at 3T. Neuroimage 2021; 241: 118430.

30. Dong Z, Milak MS, Mann JJ. (1) H MRS thermometry: impact of separately acquired full water or partially suppressed water data on quantification and measurement error. NMR Biomed 2021: e4681.

31. Verius M, Frank F, Gizewski E, Broessner G. Magnetic Resonance Spectroscopy Thermometry at 3 Tesla: Importance of Calibration Measurements. Ther Hypothermia Temp Manag 2019; 9(2): 146–155.

32. Cavassila S, Deval S, Huegen C, van Ormondt D, Graveron-Demilly D. Cramer-Rao bound expressions for parametric estimation of overlapping peaks: influence of prior knowledge. J Magn Reson 2000; 143(2): 311–320.

33. Murakami T, Ogasawara K, Yoshioka Y, Ishigaki D, Sasaki M, Kudo K et al. Brain temperature measured by using proton MR spectroscopy predicts cerebral hyperperfusion after carotid endarterectomy. Radiology 2010; 256(3): 924–931.

34. Covaciu L, Rubertsson S, Ortiz-Nieto F, Ahlstrom H, Weis J. Human brain MR spectroscopy thermometry using metabolite aqueous-solution calibrations. J Magn Reson Imaging 2010; 31(4): 807–814.

